# Nucleus-specific thalamic involvement in seizure networks differentiates neuromodulation outcomes

**DOI:** 10.64898/2026.06.27.26356691

**Authors:** Brian Ji, Peter Hadar, Birgit Frauscher, Shruti Agashe, Derek Southwell, Kassem Jaber, Behnaz Esmaeili, Shahin Hakimian, Benjamin L. Grannan, R. Mark Richardson, Sydney S. Cash, Pariya Salami

**Author notes:** Correspondence to: Pariya Salami, Massachusetts General Hospital, Thier Research Building, Suite 423, 50 Blossom Street, Boston, MA 02114, USA.

## Abstract

Closed-loop neuromodulation via responsive neurostimulation (RNS) of the thalamus has emerged as a promising therapy for drug-resistant epilepsy (DRE), particularly in patients with broad or multifocal onset. However, response to thalamic RNS is inconsistent, and there is a crucial need to identify factors that distinguish responders from non-responders. Given the heterogeneous composition of the thalamus, the specific contributions of individual thalamic nuclei during seizures may explain the variability in outcomes between patients and could potentially serve as biomarkers for guiding target selection.

We analyzed 129 seizures from 28 patients with DRE who underwent stereo-EEG monitoring with recordings of the centromedian (CM: *n* = 15) or pulvinar (PLV: *n* = 13) thalamic nuclei and were subsequently treated with RNS targeting the corresponding nucleus (CM: 11/15 [73%] responders; PLV: 7/13 [54%] responders). Patients were classified as responders (Engel class I-III) or non-responders (Engel class IV) based on reduction in seizure frequency. For each seizure, we constructed functional connectivity networks spanning seizure onset to termination and quantified the role of the thalamic nucleus by computing its total node strength. We also used an automated detection algorithm to measure the time of seizure spread to each thalamic nucleus relative to seizure onset. Connectivity and spread timing were then compared between responders and non-responders within each nucleus group.

The timing of thalamic recruitment following seizure onset did not differ significantly between responders and non-responders in either nucleus, although CM responders showed a non-significant trend toward earlier recruitment. Analysis of functional connectivity revealed nucleus-specific patterns. CM responders exhibited significantly higher thalamic node strength than non-responders during the late-seizure phase, with no significant difference at early- or middle-seizure phases. PLV responders showed significantly higher thalamic node strength during the middle-seizure phase, but there was no significant difference at early- or late-seizure phases.

These findings suggest that the degree and timing of thalamic involvement during seizures may serve as biomarkers for predicting response to thalamic RNS in DRE. CM involvement in responders was characterized by stronger connectivity that persisted through seizure termination, whereas PLV involvement in responders was reflected primarily in connectivity during seizure propagation and progression. Incorporating these nucleus-specific ictal network features into pre-surgical evaluation could improve patient selection and guide nucleus-specific targeting for thalamic RNS.

## Introduction

Epilepsy is a neurological condition that affects approximately 1% of the global population, and despite anti-seizure medications (ASMs) serving as the primary treatment approach, nearly one-third of patients remain drug resistant.^1,2^ For these individuals, surgical resection of the seizure onset zone can provide seizure freedom in a subgroup of patients. However, many patients are not candidates for resective surgery because of multifocal epilepsy or seizure foci within eloquent cortex, while others continue to experience seizures despite surgery. In such cases, intracranial neuromodulation—the delivery of electrical stimulation to targeted brain regions with the aim of modulating brain activity—offers a promising alternative that can reduce seizure frequency.^3^

Neuromodulation targeting the thalamus has emerged as an increasingly popular therapeutic approach, particularly for patients with multifocal or broadly distributed seizure onsets, as the thalamus maintains connections to widespread cortical and limbic regions which position it as a structure through which seizure activity can propagate^4,5^. Deep brain stimulation (DBS) of the thalamus has been extensively studied and has demonstrated efficacy in reducing seizure frequency in patients with drug-resistant epilepsy (DRE).^6–8^ More recently, closed-loop neuromodulation of the thalamus via responsive neurostimulation (RNS) has gained increasing attention and has demonstrated encouraging early results. However, because the thalamus comprises multiple functionally distinct nuclei, clinicians must decide which nucleus to target for each patient. Because RNS requires detection of seizure activity to deliver stimulation, presurgical stereo-EEG (sEEG) evaluations are commonly used to identify the thalamic nucleus that demonstrates early ictal involvement.^9–11^ This reliance on visual inspection of sEEG traces introduces subjectivity and may fail to capture the full extent of thalamic engagement. Despite growing clinical experience, no quantitative framework exists for predicting which patients will respond to stimulation of a given thalamic nucleus, and more importantly, even when nuclei with early ictal involvement are targeted, clinical outcomes remain variable.^12^

Multiple thalamic nuclei have been investigated as targets for neuromodulation in DRE, including the anterior (ANT), centromedian (CM), and pulvinar (PLV) nuclei. Clinical experience suggests that different nuclei may be more suitable for different patients, likely reflecting differences in the underlying seizure network rather than a universally optimal target.^13^ One potential explanation is that each nucleus exhibits distinct anatomical and functional connectivity with cortical and subcortical regions, shaping how it participates in seizure initiation and propagation. Consistent with this concept, our previous work demonstrated that seizures arising from different cortical regions exhibit distinct patterns of early thalamic recruitment, with preferential involvement of specific nuclei depending on seizure onset location.^14^ We further showed that each thalamic nucleus possesses a unique interictal connectivity profile with the cortex, and that these connectivity patterns differ between responders and non-responders to thalamic RNS.^15^ Together, these findings suggest that thalamocortical connectivity may provide a quantitative measure of a nucleus’s role within an individual’s seizure network and may ultimately help guide patient-specific target selection for neuromodulation.

If thalamocortical connectivity reflects the role of a nucleus within an individual’s seizure network, it may also provide a means of predicting therapeutic response to neuromodulation. This possibility is supported by studies demonstrating that short periods of high-frequency thalamic neurostimulation reduce cortical excitability in proportion to effective connectivity between the stimulated nucleus and the seizure network, while longer-term RNS therapy is associated with reorganization of interictal functional connectivity in patients who achieve clinical benefit.^16,17^ These findings suggest that nucleus-specific connectivity may serve as a biomarker for predicting stimulation response. Such a biomarker is particularly needed for the CM and PLV nuclei, both of which maintain widespread but distinct cortical connectivity and are increasingly used as RNS targets.^10,14,18–21^ However, the network features that distinguish responders from non-responders, and the specific roles of these nuclei in mediating therapeutic benefit, remain poorly understood.

Understanding how the CM and PLV differentially participate in seizure networks, including differences in spread timing and ictal connectivity, may reveal signatures that predict which patients will benefit from stimulation of a particular target.^22^ We therefore hypothesized that nucleus-specific seizure network connectivity patterns could serve as biomarkers of thalamic RNS response. Here, we leveraged sEEG recordings from patients with CM and PLV electrodes who subsequently underwent thalamic RNS to investigate whether nucleus-specific ictal connectivity patterns distinguish treatment responders from non-responders, and whether the timing of seizure spread to each nucleus differs in a clinically meaningful way. Our findings reveal that the CM and PLV exhibit distinct connectivity-outcome relationships, and these connectivity biomarkers could enable preoperative prediction of thalamic RNS response for individual patients, moving toward a more patient-specific framework for epilepsy neuromodulation.

## Materials and methods

### Patient selection

This was a multi-center retrospective study conducted at Duke University, the University of Washington, and Mass General Brigham. The study was approved by the institutional review board (IRB) at each participating site, and the requirement for informed consent was waived given the retrospective nature of the analysis. We identified all patients with DRE at the three participating institutions who underwent sEEG monitoring with at least one electrode sampling the CM or PLV thalamic nucleus and were subsequently implanted with an RNS system targeting the same nucleus that was recorded during sEEG. Both adult and pediatric patients were included. Patients were excluded if post-RNS follow-up was less than 6 months, as this duration was required for reliable outcome assessment. After applying these criteria, 28 patients were included in the final cohort, comprising 15 patients in the CM group (11 responders, 4 non-responders) and 13 patients in the PLV group (7 responders, 6 non-responders), with a total of 129 analyzed seizures.

### Electrode implantation and recording

Electrode selection and trajectory planning were performed independently at each participating institution by the local epilepsy team based on the clinical hypothesis of the seizure onset zone, without input from the present study. Stereotactic depth electrodes were implanted, with at least one electrode trajectory designed to sample the CM or PLV thalamic nucleus in addition to the cortical and subcortical regions of clinical interest. Intracranial recordings were acquired using the Blackrock system with FrontEnd amplifiers (Blackrock Microsystems), Pegasus Acquisition Software (Neuralynx), or Natus NeuroWorks system (Natus Medical Incorporated) at a sampling rate of 1024Hz or 2048Hz and extracted for analysis.

Electrode contacts were localized using a modified version of the pipeline described by Soper et al.^23^ For each patient, a pre-implantation T1- and T2-weighted MRI was co-registered with a post-implantation CT scan in patient-native space. A full cortical and subcortical reconstruction of the pre-implantation MRI was performed using FreeSurfer’s recon-all pipeline.^24^ Individual electrode contact coordinates were extracted from the co-registered CT and manually verified on the patient’s native MRI.

Thalamic nuclei were segmented using FreeSurfer’s probabilistic thalamic nuclei segmentation.^25^ Electrode contacts were then assigned to specific thalamic nuclei using the Electrode Volume Labeling (EVL) approach.^23^ The thalamic segmentation volumes were exported and imported into MATLAB (R2024b; MathWorks, Natick, MA), and each electrode contact was assigned to a thalamic nucleus if its coordinates fell within the corresponding volume. The two target nuclei of interest were the centromedian and the pulvinar nuclei, corresponding to the CM and PuM labels in the FreeSurfer thalamic atlas, respectively. For bipolar contact pairs, a pair was classified as being within a given nucleus if at least one of the two contacts was contained within that nucleus.

To illustrate the locations of these electrodes, we mapped the electrode locations to common brain locations in MNI (Montreal Neurological Institute) space using MATLAB and FieldTrip tools.^26^ The sEEG and RNS electrodes targeting the CM and PLV were visualized using Lead-DBS with the Morel Atlas.^27,28^

### Seizure selection and annotation

For each patient, seizures recorded during sEEG monitoring were reviewed and selected for analysis based on the following criteria. Seizures with focal onset (i.e., onset restricted to a single contact or region) that remained focal throughout the length of the seizure were excluded, as the current analysis focused on the role of thalamic nuclei in seizures with broader or multifocal onset patterns. Among the remaining non-focal seizures, distinct seizure types were defined for each patient based on the combination of clinical semiology and electrographic onset location. To prevent any single seizure type or patient from dominating the analysis, a maximum of five seizures per type per patient were included; when more than five seizures of a given type were available, the most representative and electrographically clearest examples were selected.

Seizure onset, termination, and channel-level involvement were annotated by the clinical team at each participating institution and subsequently verified by a board-certified epileptologist. Seizure onset was defined as the earliest unequivocal electrographic change from the immediately preceding background activity. Seizure termination was defined as the time point at which sustained ictal activity ceased, regardless of whether it was followed by postictal suppression, slowing, or return to baseline activity.

### RNS outcome assessment

Outcomes following RNS implantation were assessed clinically and used to classify patients as responders or non-responders. Follow-up duration was measured from the date of RNS activation to the most recent clinic visit, with a minimum required duration of 6 months. Post-implantation seizure frequency was quantified from patient-maintained seizure diaries and compared against pre-implantation baseline, defined as the mean monthly seizure frequency in the months immediately preceding RNS activation.

Treatment response was graded using the Engel outcome scale based on the percent reduction in disabling seizures at the most recent follow-up visit: Engel class I, free from disabling seizures; Engel class II, rare disabling seizures (almost seizure free); Engel class III, worthwhile improvement; and Engel class IV, no worthwhile improvement. Patients with Engel class I–III outcomes were classified as responders, and patients with Engel class IV outcomes were classified as non-responders. Outcome assessment was performed by a clinician blinded to all sEEG connectivity and seizure-spread analyses.

### Seizure spread timing analysis

To quantify when ictal activity spreads to each thalamic nucleus of interest, we applied an automated detection algorithm adapted from Salami et al.^14^ Seizures with duration shorter than 15 seconds or longer than 300 seconds were excluded from this analysis. For each remaining seizure, a single bipolar pair representative of each recorded thalamic nucleus was selected per hemisphere. When both ipsilateral and contralateral thalamic recordings were available, the side ipsilateral to the seizure onset was used. For seizures with broad bilateral onset, the earliest detection across the two hemispheres was retained as the time of spread.

For each thalamic channel, seven signal features were computed in 2-second sliding windows with a 1-second step (50% overlap): line length, area under the curve, standard deviation of the voltage trace, and spectral power in the theta, alpha, beta, and gamma bands. Each feature time series was normalized by subtracting the mean and dividing by the standard deviation of the feature values computed during the reference period, defined as the first 15 seconds of the epoch (−30 s to −15 s relative to onset). A window was considered significantly elevated if its normalized value exceeded twice the standard deviation of the reference period. For each feature, a seizure was determined to have spread to the channel if the feature was significantly elevated for at least three consecutive windows within the 30 seconds following seizure onset; the time of spread for that feature was defined as the first of these three consecutive windows. The earliest detection across the seven features was taken as the time of spread for that channel.

To determine which electrographic features detected thalamic recruitment earliest and most reliably within each nucleus, we extended the spread-timing analysis to characterize each of the seven signal features individually. For this analysis, seizures were grouped by nucleus (CM and PLV) and pooled across responders and non-responders. For each feature, we computed two quantities across the cohort. The detection rate was defined as the proportion of seizures in which that feature produced a sustained significant elevation (at least three consecutive windows exceeding the reference threshold) within the 30 seconds following seizure onset. Among seizures in which the feature reached this criterion, the detection latency was reported as the median [interquartile range] time from seizure onset. Features were then ranked by median detection latency to identify the earliest-detecting feature in each nucleus.

### Connectivity analysis

For each seizure, time-resolved functional connectivity networks were constructed using the nonlinear correlation coefficient h² as implemented in AnyWave.^29^ Pairwise directed h² matrices were computed between all bipolar channels using sliding windows of 2 seconds with a 1 second step, separately for broadband (1-55 Hz) and five component frequency bands: delta (1-4 Hz), theta (4-8 Hz), alpha (8-12 Hz), low beta (12-20 Hz), and high beta (20 – 30 Hz). For each window, this yielded an nCh × nCh directed adjacency matrix in which entry (i, j) reflects the strength of nonlinear association from channel i to channel j.

To quantify the degree of thalamic involvement in each seizure network, we computed the total node strength of each thalamic channel at every window. Total node strength was defined as the sum of in-strength (the column sum of the adjacency matrix) and out-strength (the row sum), capturing both incoming and outgoing connectivity. For each seizure, only the thalamic side that showed the earliest spread (see Seizure spread timing analysis) was retained; when contacts from both hemispheres of the same nucleus were available for that side, node strength was averaged across them.

Connectivity matrices were computed for four temporal periods relative to each seizure: a pre-ictal baseline (−30 to −15 seconds before onset), the ictal period (seizure onset to nucleus-specific termination), and pre-seizure and post-seizure peri-ictal windows, computed at the same window and step size. To allow group-level averaging across seizures of varying duration, each ictal node-strength trace was linearly interpolated to a fixed length of 100 windows spanning seizure onset to termination. Each interpolated trace was then baseline-normalized by subtracting the per-channel mean of the pre-ictal baseline period.

### Statistical analysis

Statistical analyses were performed in Python 3.12 with SciPy and NumPy, and MATLAB R2024b (MathWorks, Natick, MA).^30,31^ For the seizure spread timing analysis, within each thalamic nucleus group (CM and PLV), spread times were compared between responders and non-responders using two complementary tests: a Wilcoxon rank-sum (Mann–Whitney U) test on the continuous spread times, and a Fisher’s exact test on the contingency of early versus late spread. Statistical significance was set at p < 0.05 for both tests. To test whether detection latency differed across features, we used a Friedman test across all seven features, treating the individual seizure as a paired observation. Seizures in which a given feature never met the detection criterion were assigned a latency equal to the 30-second post-onset analysis window for the purpose of this test. This treated non-detections as maximally late. Post hoc comparisons against the earliest-detecting feature were performed using Wilcoxon signed-rank tests, and statistical significance was set at p < 0.05. For the connectivity analysis, to compare responders and non-responders, we used a seizure-level bootstrap with 10,000 resamples. At each iteration, seizures were sampled with replacement separately within the responder and non-responder groups, and the mean node-strength trace was computed for each group; the difference between group means was then evaluated at every time point. Two-sided p-values were derived empirically from the distribution of bootstrap differences, and time points with p < 0.05 were considered significant; no multiple-comparison correction was applied across the 100 time points. The 100-window ictal trace was divided into three equal phases corresponding to the early-seizure (windows 1–33), mid-seizure (windows 34–66), and late-seizure (windows 67–100) phases, and between-group differences were summarized within each phase.

## Results

### Patient cohort and seizure dataset

We analyzed sEEG recordings from 28 patients with DRE who underwent thalamic sampling of the CM or PLV nucleus and were subsequently treated with RNS targeting the same nucleus. Fifteen patients had recordings from the CM and 13 from the PLV. The CM group comprised 11 responders and 4 non-responders, and the PLV group comprised 7 responders and 6 non-responders. Of 165 seizures reviewed across the 28 patients, 36 were excluded after applying the inclusion criteria (focal-onset seizures and the cap of five seizures per type per patient), yielding 129 seizures for analysis (76 CM, 53 PLV). Representative sEEG and RNS electrode reconstructions are shown for both nuclei in Figure 1 (Fig. 1A-D).

**Figure 1.**
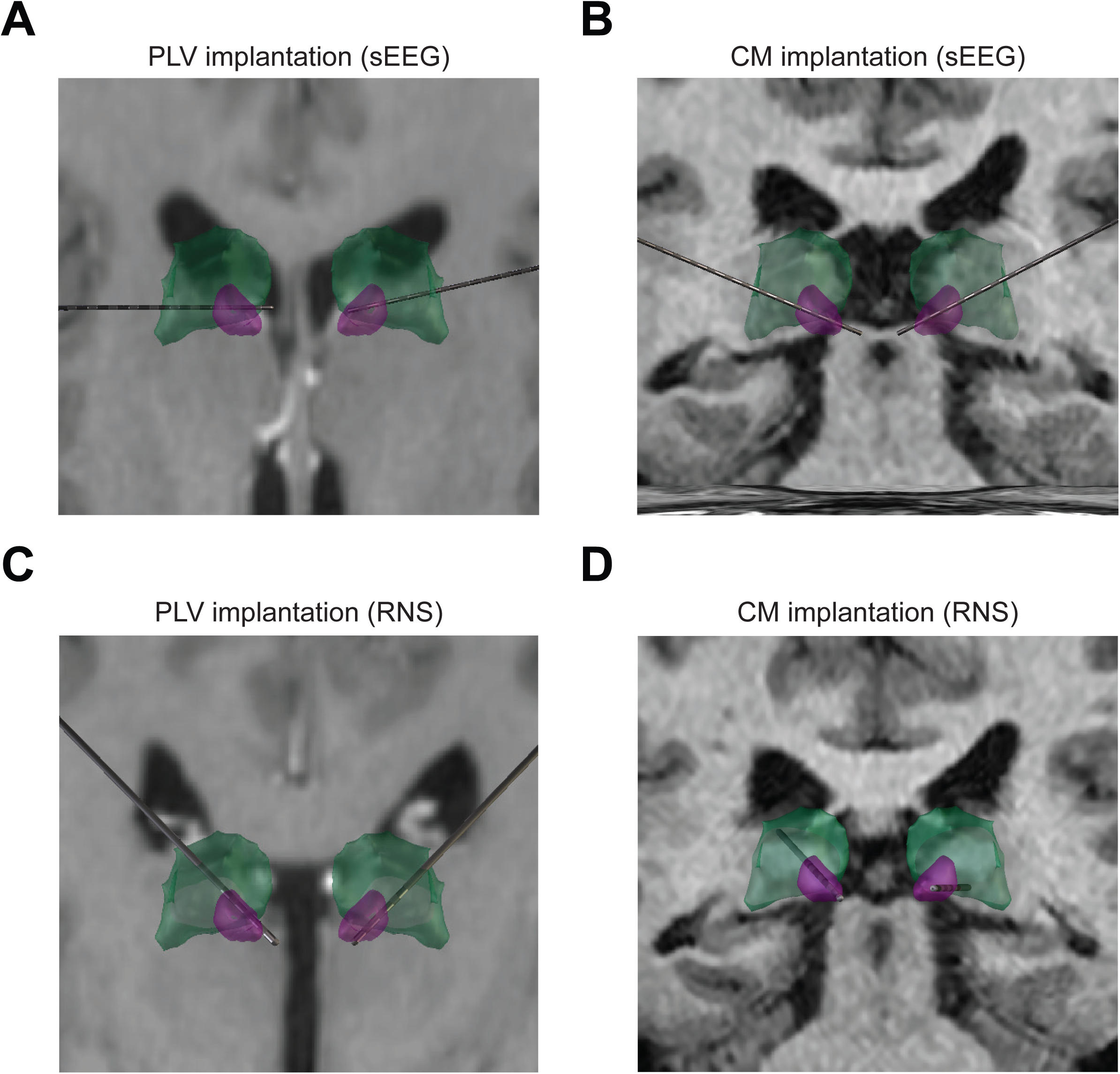
Reconstructions of sEEG and RNS electrodes and example seizures captured from sEEG. (A) Example reconstruction of sEEG electrodes in a patient with bilateral pulvinar contacts. (B) Example reconstruction of sEEG electrodes in a patient with bilateral centromedian and pulvinar contacts. (C) Example reconstruction of RNS electrodes in a patient with bilateral pulvinar RNS contacts. (D) Example reconstruction of RNS electrodes in a patient with bilateral centromedian RNS contacts.

### Timing of thalamic recruitment did not distinguish responders from non-responders

We first asked whether the latency of seizure spread to each thalamic nucleus differed between responders and non-responders. In the CM cohort, spread times did not differ significantly between groups: responder seizures reached the CM at a median of 1.0 s after seizure onset (57 seizures, 11 patients), compared with 2.0 s for non-responder seizures (19 seizures, 4 patients; Wilcoxon rank-sum p = 0.27). Dividing seizures into early (Σ6 s) versus late spread yielded a similar null result (Fisher’s exact p = 0.40) (Fig. 2E). In the PLV cohort, spread times were likewise indistinguishable between groups, with a median of 1.0 s in both responder seizures (29 seizures, 7 patients) and non-responder seizures (24 seizures, 6 patients; rank-sum p = 0.29; Fisher’s exact p = 1.0) (Fig. 2F). Therefore, the latency of seizure spread to either thalamic nucleus did not distinguish responders from non-responders, indicating that spread timing alone is insufficient to predict RNS responsiveness and motivating the connectivity-based analyses described below.

**Figure 2.**
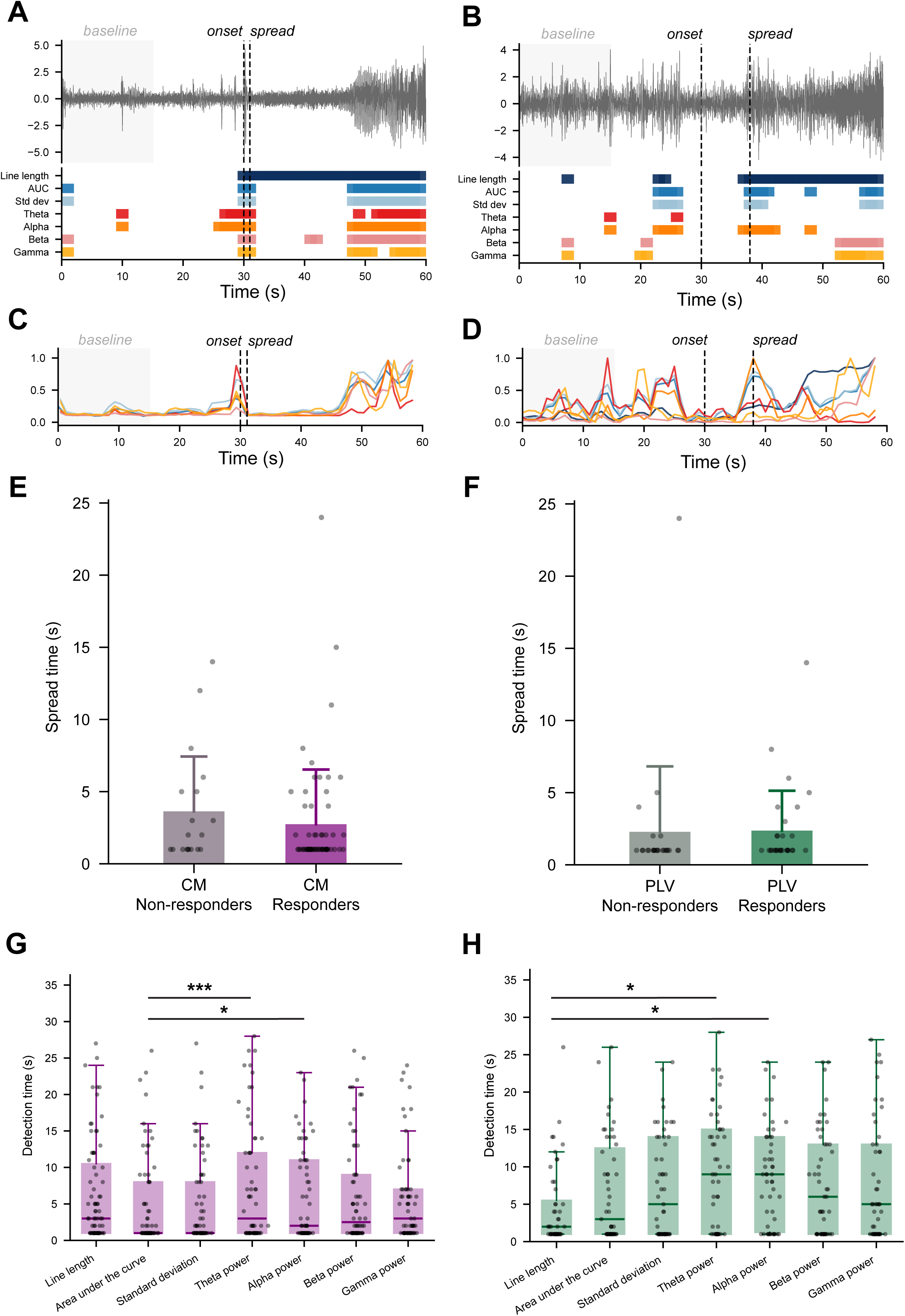
Thalamic spread timing and correlation with RNS responder status. Example results from two seizures showing signal traces with corresponding feature significance maps (A, B) and normalized feature time courses (C, D). In the left example (A, C), spread is detected shortly after onset, whereas in the right example (B, D), spread is detected with a longer delay. (E) Distribution of seizure spread times to the CM for responders (purple) and non-responders (gray), with each point representing a single seizure. (F) Distribution of seizure spread times to the PLV for responders (green) and non-responders (gray). (G) Distribution of detection latencies for each feature among seizures in which the feature fired, shown as box plots with seizures for CM (purple). (H) Distribution of detection latencies for each feature among seizures in which the feature fired, shown as box plots with seizures for PLV (green). Asterisks denote features detecting significantly later than the earliest-detecting feature within each nucleus (Wilcoxon signed-rank, p < 0.05).

### Feature-specific detection of thalamic recruitment

Across the CM cohort (76 seizures, 15 patients), all seven features detected thalamic recruitment in the majority of seizures, with detection rates ranging from 75.0% (theta, alpha, and gamma power, 57/76) to 90.8% (line length, 69/76). The earliest detections were driven by amplitude-and energy-based features: area under the curve and standard deviation each reached criterion at a median of 1.0 s (IQR 1.0-9.0) after seizure onset, with area under the curve serving as the earliest-detecting reference feature. Line length, despite having the highest detection rate, detected later (median 4.0 s [1.0-11.0]), and the spectral power features detected at intermediate latencies (alpha power, 2.0 s; beta power, 3.5 s; gamma power, 5.0 s; theta power, 6.0 s). Detection latency differed significantly across features (Friedman = 25.3, p = 0.0003, n = 76), and post hoc comparisons showed that area under the curve detected significantly earlier than theta power (p = 0.0014) and alpha power (p = 0.028), but not significantly earlier than line length, standard deviation, beta power, or gamma power (all p > 0.09). Detection latencies in CM were thus short and clustered, with all median latencies falling between 1.0 and 6.0 s (Fig. 2G, H).

In the PLV cohort (53 seizures, 13 patients), detection rates were uniformly high (86.8-92.5%), with area under the curve and standard deviation detecting most reliably (92.5%, 49/53 each). In contrast to CM, line length was the earliest-detecting feature (median 2.0 s [1.0-6.5]), and detection latencies spanned a substantially wider range across features. Spectral power measures detected later, with theta and alpha power both reaching criterion at a median of 9.0 s (IQR 1.0-15.0 and 1.75-14.0, respectively), while standard deviation, beta power, and gamma power detected at intermediate latencies (5.0, 6.0 and 5.0 s). Detection latency again differed significantly across features (Friedman = 24.4, p = 0.0004, n = 53), and post hoc comparisons showed that line length detected significantly earlier than alpha power (p = 0.049); the comparison against theta power showed a similar trend that did not reach significance (p = 0.072), and the remaining features did not differ significantly from line length (all p > 0.10).

### Broadband thalamic connectivity distinguishes responders in a nucleus- and phase-specific manner

We next compared the strength of thalamic connectivity, quantified as broadband (1-55 Hz) node strength normalized to the pre-ictal baseline, between responders and non-responders across the seizure. Each seizure was time-normalized to 100 ictal windows with 10 pre- and 10 post-ictal windows. Responder and non-responder node strength was compared at each window using a bootstrap procedure, with significance defined as a 95% confidence interval of the responder-non-responder difference that excluded zero.

In the CM cohort, responder seizures (n = 58) showed significantly greater broadband node strength than non-responder seizures (n = 18) during the later phases of the seizure. The difference reached significance across a contiguous block of windows spanning the middle and final ictal thirds and extending through termination (windows 64-98 within the ictal window), with two additional brief differences in the immediate post-termination period. Node strength did not differ between groups during seizure onset and propagation (first ictal third) (Fig. 3).

**Figure 3.**
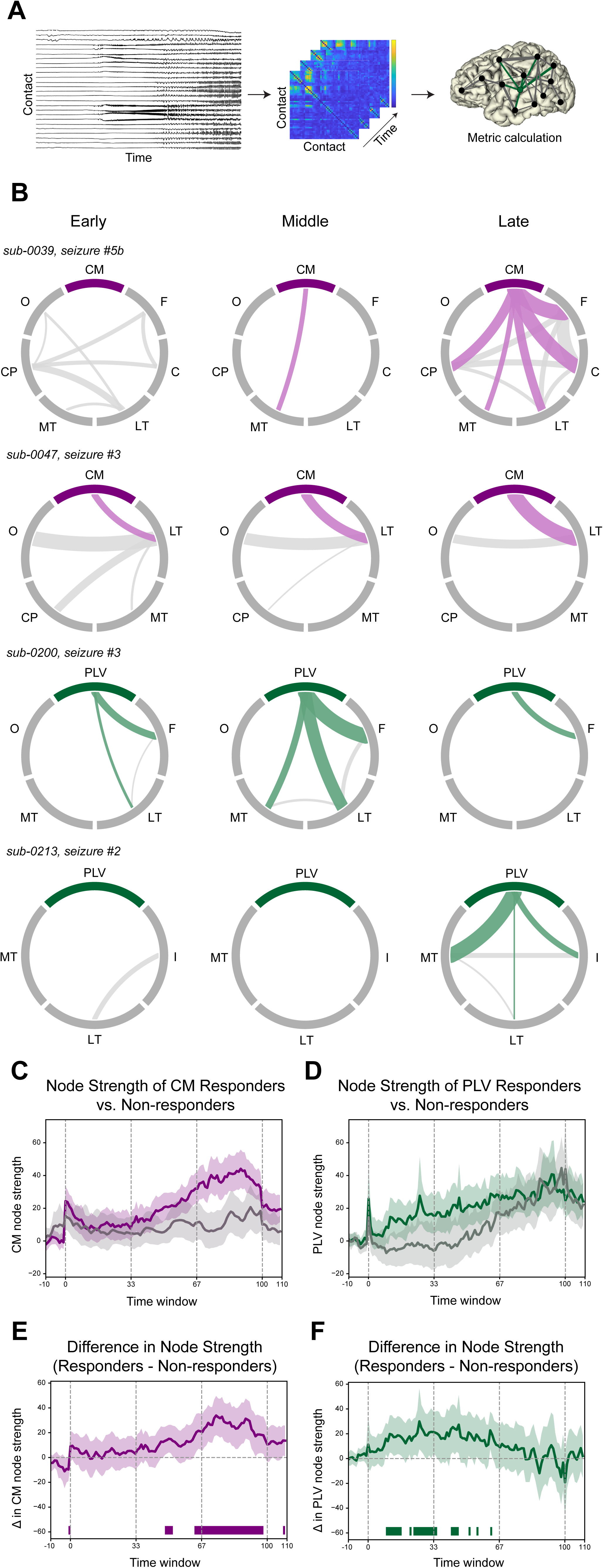
Broadband thalamic connectivity pipeline and group-level comparison of node strength between RNS responders and non-responders. (A) Analysis pipeline schematic: bipolar sEEG recordings are used to compute h^2^ connectivity matrices across sliding windows, from which graph-theoretic metrics are extracted at thalamic contacts. (B) Example circular connectivity plots of seizures from a responder to CM RNS (sub-0039), non-responder to CM RNS (sub-0047), responder to PLV RNS (sub-0200), and non-responder to PLV RNS (sub-0213); F: frontal, C: cingulate, LT: lateral temporal, MT: mesial temporal, CP: centroparietal, O: occipital, I: insular (C, D) Broadband node strength timecourses at the CM (purple) and PLV (green) nuclei for responders (colored) and non-responders (gray), shown as bootstrapped means with 95% confidence intervals. (E, F) Bootstrapped difference in node strength (responders minus non-responders) for CM and PLV, with colored bars indicating statistically significant time windows (p < 0.05, 10000 iterations). Dashed lines delineate first third (early phase), second third (middle phase), and last third (late phase).

In the PLV cohort, the responder-nonresponder difference emerged earlier in the seizure. Responder seizures (n = 27) showed significantly greater broadband node strength than non-responder seizures (n = 26) across multiple windows spanning the first and second ictal thirds (windows 9-63), with no significant difference during termination or the post-ictal period (Fig. 3). To complement the window-level analysis, we summarized broadband node strength within five phases (pre-ictal; early [T1], middle [T2], and late [T3] ictal thirds; and post-ictal) and compared responders with non-responders in each phase. This phase-level analysis reproduced the nucleus-specific temporal dissociation. In the CM cohort, responders showed significantly greater node strength than non-responders during the late-ictal phase (T3: p = 0.022), with no significant difference during the pre-ictal, early phase (T1: p = 0.66), middle phase (T2: p = 0.33), and post-ictal phase. In the PLV cohort, responders showed significantly greater node strength during the middle-ictal phase (T2: p = 0.034), with a similar trend in the early phase that did not reach significance (T1: p = 0.21) and no difference during the late-ictal (T3: p = 0.86), pre-ictal, or post-ictal phases. Notably, in the PLV, both groups reached comparably high node strength by the late-ictal phase (median 26.7 vs. 29.6), indicating that the responder-non-responder difference was confined to seizure progression rather than termination.

To illustrate these group-level differences at the single-seizure level, we visualized region-to-region thalamocortical connectivity for representative seizures across the three ictal phases (early, middle, and late; Fig. 3B). In a representative CM responder (sub-0039), CM-cortical connectivity was sparse during propagation and progression and increased by termination, with strong connections emerging from the CM to frontal, cingulate, lateral and mesial temporal, and centroparietal regions, consistent with the late-phase CM effect observed at the group level. In a representative CM non-responder (sub-0047), CM connectivity remained comparatively sparse. In contrast, in a representative PLV responder (sub-0200), PLV-cortical connectivity was most prominent during propagation and progression, mostly to lateral and mesial temporal and frontal regions, and diminished by termination. This mirrors the early-to-mid PLV effect. In a PLV non-responder example (sub-0213), thalamocortical connectivity was minimal during propagation and progression and appeared only at termination, displaying the inverse of the PLV-responder profile.

Therefore, although responders in both cohorts exhibited stronger ictal thalamic connectivity than non-responders, the phase in which this difference appeared was nucleus-specific: in the CM, it was concentrated in the late-ictal and termination phases, whereas in the PLV, it was concentrated in the early-to-mid phases. This dissociation indicates that the seizure phase during which a thalamic nucleus is most strongly engaged, rather than connectivity strength alone, characterizes the responder profile, and that the relevant phase differs between the two targets.

### Responder connectivity differences are concentrated in lower frequency bands

To determine which frequency bands carried these connectivity differences, we repeated the node-strength comparison within five frequency bands (delta, theta, alpha, low-beta, and high-beta). The responder pattern was driven primarily by the lower-frequency bands. In the delta, theta, and alpha bands, responders again showed significantly greater node strength than non-responders, and the nucleus-specific phasing was preserved: CM differences were concentrated in the middle and late ictal phases (for example, delta windows 74-102 and theta windows 77-103), whereas PLV differences were concentrated in the early-to-mid phases (for example, delta windows 25-72 and theta windows 20-38). The largest effects occurred in the delta and theta bands, comparable in magnitude to broadband (Fig. 4).

**Figure 4.**
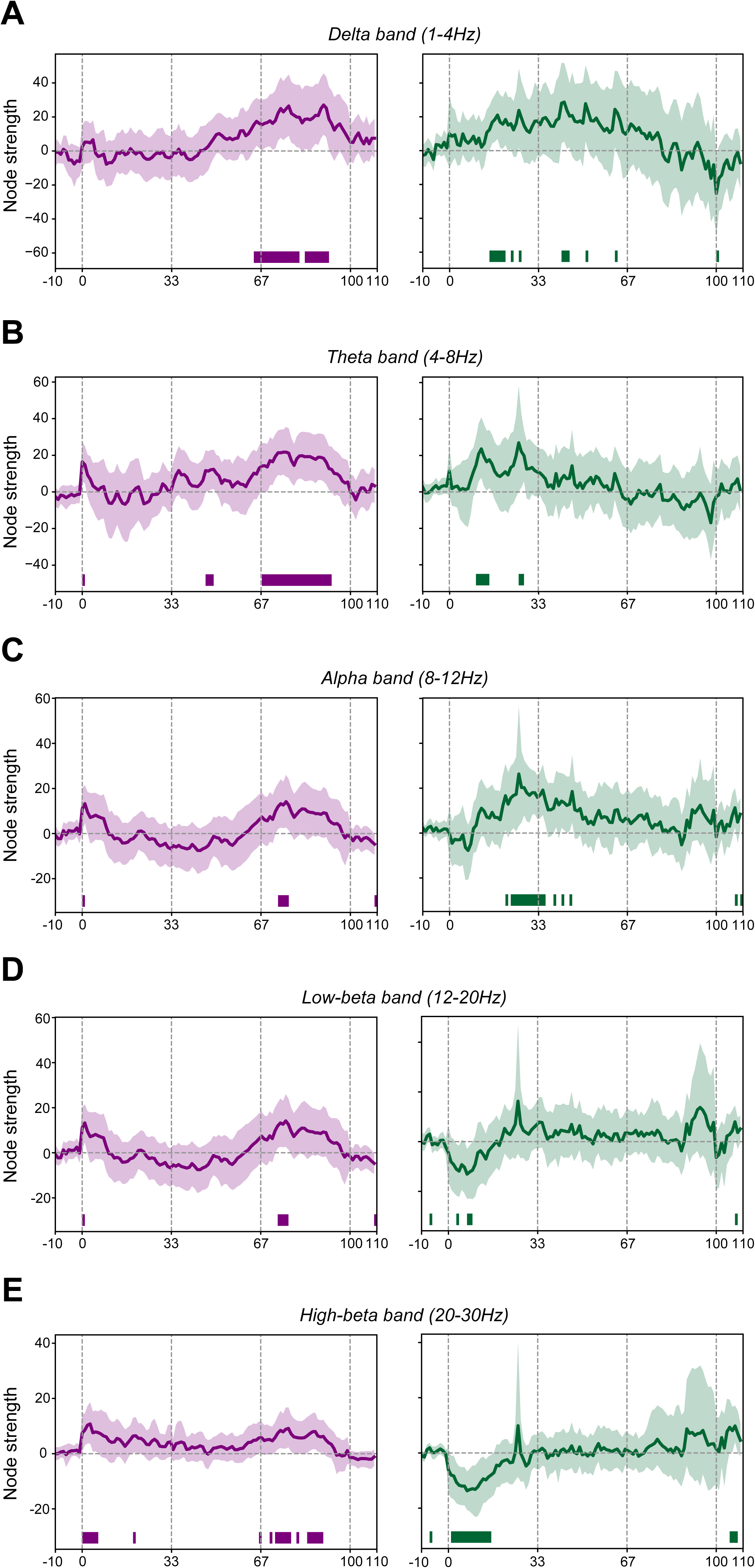
Frequency band-specific differences in thalamic node strength between RNS responders and non-responders. (A-E) Bootstrapped difference in thalamic node strength (responders minus non-responders) within five frequency bands, shown for the centromedian (CM, purple) and pulvinar (PLV, green) nuclei: (A) delta (1-4 Hz), (B) theta (4-8 Hz), (C) alpha (8-12 Hz), (D) low-beta (12-20 Hz), and (E) high-beta (20-30 Hz). For each band and nucleus, traces show the bootstrapped mean difference with 95% confidence intervals, and colored bars indicate time windows of statistically significant difference (p < 0.05, 10,000 iterations); positive deflections indicate greater node strength in responders. Dashed lines delineate the first third (early phase), second third (middle phase), and last third (late phase) of the seizure, as in Figure 3.

In the beta bands, this pattern became nucleus-specific. CM responders continued to show greater node strength than non-responders during the late ictal phase, whereas in the PLV the effect reversed: responder seizures showed significantly lower beta node strength than non-responders during the early ictal phase. The elevated thalamic connectivity that characterized responders was therefore concentrated in the lower-frequency bands, spanning delta through alpha, where it preserved the nucleus-specific phasing observed for broadband, and became CM-specific or reversed in the beta range.

Across these bands, significance was defined by bootstrap confidence intervals at individual time windows and was not corrected for multiple comparisons; we therefore base our interpretation on sustained, multi-window differences, such as the contiguous delta and theta runs, rather than on isolated single-window comparisons.

## Discussion

Thalamic neuromodulation has emerged as an increasingly important therapeutic strategy for patients with DRE. While the ANT has long been the most established and FDA-approved thalamic target for focal epilepsy, targeting the CM and PLV has expanded substantially over the past several years. CM has traditionally been favored in generalized or multifocal epilepsies, including Lennox-Gastaut syndrome and broader thalamocortical network epilepsies, whereas PLV has increasingly been utilized in patients with posterior quadrant, temporo-parieto-occipital, or broader cortical epilepsies involving posterior network propagation. Although DBS was the first major platform for thalamic neuromodulation, RNS offers several practical and mechanistic advantages, including patient-specific seizure detection, adaptive stimulation, and the ability to record chronic ambulatory electrographic data^32–34^.

At present, selection of CM or PLV as an RNS target for poorly localized or focal epilepsies is largely guided by sEEG where clinicians evaluate whether seizure activity spreads early into a candidate nucleus and whether this activity can provide a reliable biomarker for responsive detection and stimulation. In practice, earlier recruitment of a thalamic nucleus is often interpreted as evidence that the nucleus is sufficiently embedded within the seizure network to serve as an effective intervention point. However, despite this rationale, clinical response remains heterogeneous, and not all patients with early thalamic involvement derive equivalent benefit from stimulation. This discrepancy suggests that simple timing of seizure spread may not fully capture the therapeutic relevance of a thalamic target. Rather, the broader functional role of the nucleus within the early-, mid-, and late-seizure phases may be more informative.

In this study, we leveraged seizure recordings from patients who subsequently underwent CM or PLV RNS implantation to determine whether ictal thalamic dynamics differ between responders and non-responders. By characterizing connectivity strength and temporal evolution of these nuclei from seizure onset through termination, we sought to identify electrophysiologic features beyond simple spread latency that may better predict therapeutic responsiveness.

The principal findings of this study are: (1) Responders and non-responders demonstrated distinct thalamic connectivity profiles, with these differences being nucleus- and seizure-phase specific: in PLV responders, stronger connectivity was observed primarily earlier during the seizure, whereas in CM responders, connectivity differences were most pronounced during later seizure phases, particularly near termination; (2) Time to seizure spread into CM and PLV did not significantly differ between responders and non-responders, suggesting that spread latency alone may be insufficient for target optimization; (3) The electrophysiologic features most sensitive for seizure detection differed by nucleus, with amplitude- and energy-based features such as area under the curve providing the earliest detection in CM and line length providing the earliest detection in PLV.

Our findings challenge the sufficiency of prioritizing solely on earliest thalamic spread when selecting between nuclei during sEEG evaluation. We did not observe major differences in spread timing between responders and non-responders, suggesting that early spread alone may identify network participation but not necessarily therapeutic leverage. This emphasizes the need to move beyond binary “early vs late” frameworks toward richer network-based biomarkers incorporating connectivity strength, temporal phase specificity, and spectral signatures. We began to investigate whether there were nucleus-specific detection signatures. Recruitment of the CM was identified earliest by broadband amplitude and energy features, with little variation across features, whereas recruitment of the PLV was identified earliest by line length and showed wider variation, with spectral power measures lagging by several seconds. In CM, the earliest feature detected spread significantly quicker than theta and alpha power, whereas in PLV it detected spread significantly quicker than alpha power. In both nuclei, the low-frequency spectral measures displayed the latest median detections, indicating that these were consistently slower to register thalamic recruitment regardless of nucleus.

These temporal features may have important implications for closed-loop neuromodulation, as features associated with early thalamic recruitment could help identify opportunities for early intervention, whereas features emerging later in the seizure may reflect stages at which modulation of the established network is more effective. In other words, they may inform not only where to stimulate, but also when stimulation is most likely to disrupt seizure propagation.

A central contribution of this work is the demonstration that thalamic connectivity strength, rather than the mere presence or timing of seizure spread, differentiates responders from non-responders. Across both nuclei, patients who responded favorably to treatment generally exhibited stronger thalamic integration within the seizure network, suggesting that therapeutic responsiveness may depend more on how a nucleus participates dynamically within epileptic circuits than simply whether it is recruited. Importantly, this effect was temporally distinct between nuclei.

Several mechanisms may explain why stronger thalamic engagement is associated with favorable response. Node strength indexes how extensively a thalamic contact is coupled to the rest of the ictal network, so a nucleus with greater node strength is more deeply embedded as a hub within the seizure network. From the perspective of network control theory, the influence a stimulation site can exert over evolving brain dynamics depends on its connectivity to the wider network, and the energy required to steer the network away from an ictal state varies across the early, middle, and late phases of a seizure^35^. A thalamic target that is strongly, and phase-appropriately, coupled to the seizure network is therefore better positioned to propagate the effects of responsive stimulation through that network, whereas a weakly coupled nucleus may be recruited by the seizure without being able to shape it. Consistent with this framework, broader pre-treatment network connectivity beyond the seizure onset zone has been shown to predict RNS response^36,37^. Greater ictal thalamic connectivity in responders may thus reflect a nucleus that is functionally positioned to act as an effective control point within the individual patient’s seizure network.

In PLV, increased connectivity was greatest during seizure propagation and early progression, suggesting that PLV may be more functionally relevant during early network recruitment and seizure spread. This supports the notion that PLV may function as an early network amplifier or synchronizer in specific epilepsies. In contrast, CM connectivity differences were most prominent later in seizures, particularly near seizure termination, suggesting that CM may play a greater role in late-stage synchronization or seizure maintenance of widespread corticothalamic recruitment. Together, these findings suggest that CM and PLV are unlikely to represent interchangeable thalamic nodes. Rather, their differential engagement reflects distinct therapeutic entry points into different temporal phases of seizure evolution and positions within the seizure network. These putative roles align with the nuclei’s known network properties. The PLV maintains extensive reciprocal connections with parietal, occipital, and temporal cortices that position it to participate in early posterior seizure propagation^19^, whereas the CM is embedded within diffuse thalamocortical circuits implicated in generalized synchronization. Notably, cortical discharges have been shown to lead the CM during generalized epileptic activity in Lennox-Gastaut syndrome^38^, consistent with a role for the CM in sustaining rather than initiating widespread synchronization.

These distinctions carry important mechanistic implications. If PLV engagement reflects early propagation, PLV-targeted stimulation may be most effective when seizure detection and intervention occur rapidly enough to disrupt early network spread. Conversely, CM may be more relevant for modulating sustained or generalized thalamocortical synchronization later in seizure progression. Such nucleus-specific temporal roles may help explain why certain seizure phenotypes or network architectures preferentially respond to one thalamic target over another despite similar spread timing. They also suggest that the therapeutic efficacy of closed-loop neuromodulation may depend not only on where stimulation is delivered, but also when it is delivered, with interventions timed before or during specific phases of seizure evolution potentially providing greater disruption of pathological network activity.

Our frequency-specific findings further reinforce this interpretation by suggesting that responder-associated connectivity differences are driven predominantly by lower-frequency network dynamics rather than exclusively by higher-frequency pathological activity. This observation is particularly notable in the context of growing evidence that therapeutic thalamic neuromodulation may be frequency dependent. In prior work by Aiello et al. (2023), patients responding to ANT DBS demonstrated higher theta (4-8 Hz) oscillatory power within the ANT and greater network-level theta synchronization, suggesting that enhanced theta-network engagement may represent a key physiological marker of therapeutic responsiveness.^39^ Similarly, our previous work demonstrated that response to CM and PLV stimulation is associated with distinct regional and frequency-specific connectivity signatures between thalamic targets and the seizure onset zone, indicating that different thalamic nuclei may exert therapeutic effects through separable spectral and anatomical pathways.^15^

Taken together, these converging findings across ANT, CM, and PLV suggest that thalamic neuromodulation efficacy may not be explained solely by anatomical target selection, but rather by the interaction between target location, network architecture, and frequency-specific physiological states. The prominence of lower-frequency connectivity in our current cohort raises the possibility that therapeutically relevant thalamic network modulation may depend more on large-scale synchrony and oscillatory coordination than solely on suppression of fast pathological discharges. This broader framework supports the concept that different thalamic nuclei may serve as distinct frequency-tuned network control points, each with potentially unique roles in seizure modulation depending on seizure phase and underlying epileptic network organization.

From a clinical decision-making perspective, the seizure-derived features examined here, namely connectivity strength, seizure-phase dynamics, and nucleus-specific detection signatures, could in principle be integrated into a patient-specific framework to help indicate whether an individual patient is better suited for CM versus PLV targeting. If developed and validated in larger cohorts, such models may substantially improve presurgical planning by shifting target selection from qualitative electrophysiological interpretation toward quantitative, personalized network predictions.

### Limitations

Several limitations should be considered when interpreting these findings. The cohort included patients with heterogeneous seizure types, epileptogenic zones, and underlying network architectures, all of which may influence both thalamic recruitment patterns and therapeutic response.^15,40,41^ Although we attempted to compare seizure characteristics across sEEG and chronic RNS datasets, determining which specific seizures are clinically disabling, treatment-responsive, or refractory remains inherently challenging. Chronic ambulatory RNS recordings are constrained by sparse sampling, device-specific detection thresholds, and recording truncation, which can limit direct one-to-one comparisons with seizures captured in the EMU.^17,42^ As a result, important variability in seizure phenotypes may not be fully represented in currently available datasets. Future studies will likely require more refined seizure phenotyping approaches in ambulatory settings, potentially integrating patient-reported symptom diaries, wearable physiologic monitoring, or expanded chronic recording platforms to better characterize seizure-specific treatment effects over time.

In addition, while some patients demonstrated seizure spread to both CM and PLV, the retrospective nature of this study does not allow definitive conclusions regarding whether one thalamic target would have been superior to another within the same individual. This remains a central unresolved question in thalamic neuromodulation.^21,43^ The presence of seizure involvement in multiple nuclei does not necessarily indicate equivalent therapeutic utility, and differential efficacy may reflect nucleus-specific mechanisms, broader network topology, or individualized anatomy. Prospective comparative studies will be essential to determine whether observed differences in response are driven primarily by target-specific physiology or by patient-specific seizure network organization.

Therapeutic response to thalamic RNS is also likely influenced by a broader set of anatomical and stimulation-related variables beyond the seizure-derived biomarkers examined here. While this study focused on seizure spread, connectivity strength, and temporal dynamics to predict responsiveness, the precise location of electrode implantation within CM or PLV may not fully capture clinically meaningful variation. Emerging evidence suggests that microanatomical factors—such as whether stimulation contacts are positioned within gray matter, adjacent white matter pathways, or at gray–white matter junctions—may substantially alter how stimulation engages local and distributed epileptic networks.^44,45^ Thus, even within the same nominal nucleus, subtle differences in lead placement may produce markedly different therapeutic outcomes.

Similarly, stimulation programming parameters, including frequency, amplitude, pulse width, and charge density, were not systematically incorporated into this analysis and may significantly shape treatment efficacy.^46,47^ It remains unclear whether low-versus high-frequency stimulation differentially modulates CM and PLV networks, whether certain amplitudes preferentially optimize seizure control, or how these parameters interact with patient-specific anatomy and electrophysiology. These variables likely contribute substantially to clinical response but are difficult to disentangle retrospectively.

The temporal context of stimulation may represent another important but underexplored dimension. Increasing evidence suggests that neuromodulation efficacy may depend not only on where stimulation is delivered, but also when it is delivered relative to evolving seizure risk states.^48^ For instance, Anderson et al. indicate that stimulation applied during lower-risk periods may provide more effective long-term network control than stimulation delivered during high-risk periods.^49^ Because our analyses were not designed to assess adaptive state-dependent stimulation timing, these factors could not be systematically evaluated.

Taken together, these limitations underscore that response to thalamic RNS likely reflects a multidimensional interaction among seizure network architecture, target anatomy, microstructural lead location, stimulation parameters, and temporal delivery strategy. Future work should aim to integrate these dimensions into predictive frameworks that move beyond nucleus-level targeting alone. Despite these limitations, our findings provide early evidence that optimizing thalamic RNS may require consideration not only of seizure spread timing, but also of connectivity strength, seizure-phase dynamics, and nucleus-specific electrophysiologic features, advancing the field toward a more comprehensive model of precision neuromodulation.

## Conclusion

Overall, this study provides evidence that successful therapeutic response to CM and PLV RNS is likely determined less by simple early seizure recruitment and more by the dynamic functional role each nucleus plays within the evolving seizure network. Rather than serving as interchangeable thalamic targets, PLV and CM appear to represent distinct network control nodes with differential contributions to seizure initiation, propagation, maintenance, and termination. These findings challenge conventional target-selection paradigms that prioritize spread timing alone and instead support a more sophisticated network-based framework that incorporates temporal dynamics and connectivity strength. Integrating these mechanistic distinctions into presurgical evaluation has the potential to substantially refine patient-specific target selection, improve neuromodulatory efficacy, and move the field closer to truly precision-guided thalamic neuromodulation.

## Data Availability

All data produced in the present study are available upon reasonable request to the authors.

## Acknowledgments

We are sincerely grateful to the patients who participated in this study and consented to the use of their data to improve care for future epilepsy patients. We also thank the clinical and administrative staff who facilitated data collection. This study was supported by the National Institutes of Health (1S10OD034414).

